# Improving Survival Prediction from Surrogate Endpoints Through Integration of Patient-Reported Outcomes Using Machine Learning

**DOI:** 10.1101/2025.10.02.25337154

**Authors:** Yimeng Sun, Yixuan Wang, Hoang Pham Khanh Luu, Audrey Liu, Congyu Zhang, Benyam Muluneh, Wenhao Zheng, Huaxiu Yao, Ethan Basch, Jiawei Zhou

**Affiliations:** Division of Pharmacotherapy and Experimental Therapeutics, Eshelman School of Pharmacy, University of North Carolina at Chapel Hill, Chapel Hill, North Carolina, USA; Department of Computer Science, University of North Carolina at Chapel Hill, Chapel Hill, North Carolina, USA; School of Data Science and Society, University of North Carolina at Chapel Hill, Chapel Hill, North Carolina, USA; Lineberger Comprehensive Cancer Center, School of Medicine, University of North Carolina at Chapel Hill, Chapel Hill, North Carolina, USA

## Abstract

**Introduction:** Overall survival (OS) is the gold standard for demonstrating treatment benefit in oncology but requires prolonged follow-up. Surrogate efficacy endpoints, such as progression-free survival, can shorten trial duration and reduce costs but may not consistently reflect overall survival benefit. We hypothesized that integrating patient-reported outcomes (PROs) with surrogate endpoints would improve trial-level prediction of OS benefit.

**Methods:** We reviewed clinical trials supporting FDA oncology drug approvals from 2013 to 2023 that reported OS, at least one surrogate efficacy endpoint, and PROs. Machine learning models predicted OS benefit were developed using surrogate endpoints alone or surrogate endpoint features combined with PROs. Both models were validated using repeated five-fold trial-grouped cross-validation and leave-one-trial-out validation.

**Results:** A total of 100 clinical trials (103 between-arm comparisons, 68,987 participants) met inclusion criteria. Compared with control arms, 68% of active treatment arms demonstrated OS benefit, 83.5% showed surrogate endpoints improvement, and 35.9% showed improvement in PROs. Adding PROs to machine learning models increased area under the receiver operating characteristic curve (AUC-ROC) from 0.67 (95% CI, 0.55–0.78) to 0.79 (95% CI, 0.69–0.87), an absolute increase of 0.12 indicating improved model discrimination. Leave-one-trial-out validation confirmed the improvement (AUC-ROC, 0.663 versus 0.795; paired difference, 0.132; 95% CI, 0.017–0.248).

**Conclusions:** Integrating PROs with surrogate endpoints improved prediction of OS benefit, supporting machine learning as a quantitative framework for combining complementary endpoints in oncology drug development.

## Introduction

Overall survival (OS) remains the gold standard for evaluating cancer therapies efficacy and safety, but it requires long follow-up time.(1) This not only makes clinical trials lengthy and costly, but also renders OS impractical in oncology settings such as indolent diseases or highly efficacious therapeutics where patients live for many years.(2) Surrogate efficacy endpoints like progression-free survival (PFS) and overall response rate (ORR) can be measured earlier and are widely used to guide trial evaluations and clinical decisions.(3,4) However, these alternatives often show limited correlation with OS and may fail to capture meaningful survival benefit.(5) The recent Food and Drug Administration (FDA) draft guidance reaffirmed OS as the primary regulatory endpoint, highlighting an unmet clinical need: the lack of robust surrogate markers that can reliably predict OS earlier in the trial process.(6)

Patient-reported outcomes (PROs) have strong potential to fill this gap. PROs are direct reports from patients describing their symptoms, functioning, and quality of life (QoL).(7,8) Unlike radiographic or biomarker-based measures, PROs provide a unique “host perspective” that complements traditional indicators of disease progression or treatment toxicity.(9,10) These measures are also noninvasive, low-cost, and can be collected frequently, enabling more granular assessment of changes in clinical status over time.(11,12) Extensive evidence shows that integrating PROs into oncology care improves symptom control, reduces emergency visits, and may provide earlier signals of cancer progression.(13,14)

In this study, we hypothesized that integrating PROs with surrogate endpoints could improve prediction of OS benefit in randomized oncology trials. We systematically reviewed randomized clinical trials that supported FDA oncology drug approvals from 2013 to 2023 and examined relationships among surrogate endpoints, PROs, and OS benefit in the clinical trial. We first applied logistic regression to evaluate their associations, followed by machine-learning methods to capture potentially complex and nonlinear relationships among endpoints. We then assessed whether incorporating PROs improved prediction of OS benefit beyond surrogate endpoints alone. By integrating conventional surrogate efficacy endpoints with patients’ reports of symptoms, functioning, and quality of life, this framework provides a more comprehensive and patient-centered approach to evaluating treatment benefit.

## Methods

### Data Inclusion and Exclusion

We reviewed the FDA New Drug Registry at Drugs@FDA for oncology indications approved between January 2013 and December 2023, including indications that might be subsequently withdrawn. Trials were eligible if they reported (1) an OS analysis with an explicit conclusion as to whether the between-group difference was statistically significant, (2) at least one surrogate efficacy endpoint, and (3) at least one PRO endpoint. The study selection process is summarized in **Figure 1**.

**Figure 1.**
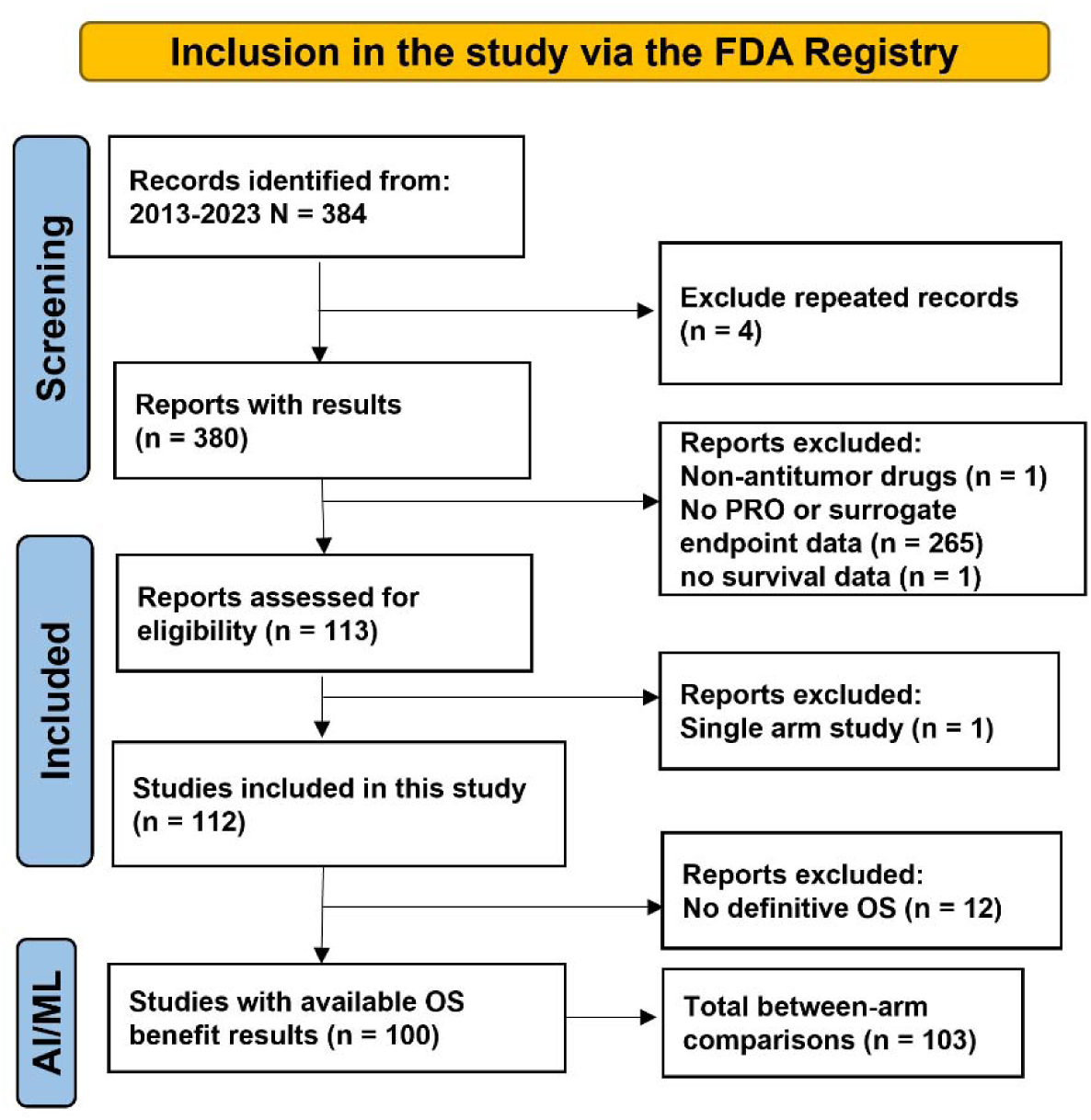
Trial inclusion and exclusion flow diagram. Randomized clinical trials of oncology drugs were identified from FDA records. Trials were included if they reported definitive OS results, at least one surrogate endpoint, and patient-reported outcomes (PROs).

### Endpoint Data Harmonization

Clinical trial data were extracted from ClinicalTrials.gov and PubMed, including endpoint measurements for active and control arms, point estimates, measures of variability, sample sizes, and p values. Because endpoint scales and directions varied across trials, all numeric treatment effects were harmonized so that positive direction-adjusted values consistently favored the active treatment. When multiple PRO endpoints were reported, global health status or overall quality of life was prioritized. If neither was available, a total score, functioning domain, or symptom endpoint that best reflected the disease- or treatment-related burden was selected. For the selected endpoints, global health status, quality-of-life, functioning, and total scores were coded as higher-is-better; symptom scores were coded as lower-is-better; and longer time to deterioration or worsening was coded as favorable. For other efficacy endpoints, longer progression-free, disease-free, event-free, or recurrence-free survival and higher event-free probabilities were coded as favorable. Hazard ratios were transformed using a negative log ratio so that values below 1 favored the active arm, and event-incidence measures were oriented so that lower incidence was favorable. For active- and control-arm point estimates, the primary continuous effect representation was an oriented symmetric difference, which places measurements reported on different scales on a comparable, benefit-oriented scale. Direction assignments and transformations were recorded in an audit table and reviewed for consistency; no included numeric endpoint had unresolved directionality. One internally inconsistent duplicate secondary PRO representation was excluded while retaining the coherent comparison record. Original trial reports were reviewed by two independent researchers to verify the reported OS-benefit classification.

### Logistic Regression Examined Associations Among Endpoints

Univariate logistic regression was used to evaluate associations of PRO and surrogate endpoint characteristics with binary OS benefit. Predictor variables included drug class, treatment benefit status, between-arm differences in endpoint point estimates, and measures of variability.

Unadjusted odds ratios (ORs) with 95% confidence intervals (CIs) were reported, and two-sided P values <0.05 were considered statistically significant.

### Machine Learning Models to Predict OS Benefits

The machine learning model prediction target was binary OS benefit. OS measurements, OS p values, and all OS-derived variables were excluded from the predictor set. Two feature sets were compared: surrogate efficacy endpoint features alone and surrogate efficacy endpoints combined with PRO features. Candidate algorithms evaluated during exploratory benchmarking included ridge and elastic-net logistic regression, Extra Trees, XGBoost, LightGBM, and a radial-basis support vector machine. Engineered predictors included direction-adjusted treatment effects between active and control arms, statistical significance based on p-values, endpoint variance, sample size and sample size ratio between treatment arms, binary indicators of endpoint benefit, endpoint characteristics, and prespecified interactions between surrogate and PRO evidence.

To prevent information leakage from multiple comparisons within the same trial, all preprocessing and validation were performed at the trial level. Numeric missing values were imputed using the training-fold median with missingness indicators and were standardized within the training fold. Categorical missing values were imputed using the training-fold mode and one-hot encoded, with infrequent categories grouped. The primary validation used 20 repeats of five-fold stratified group cross-validation, with every comparison from the same trial assigned to the same fold. Each trial contributed total weight 1, shared across its comparisons. Leave-one-trial-out validation was used as a complementary confirmation of model performance. Sensitivity analyses removing investigator-assigned indicators of endpoint benefit and sample-size-related predictors were performed to assess model robustness.

Model discrimination was primarily evaluated using the area under the receiver operating characteristic curve (AUC-ROC). For repeated cross-validation, held-out probabilities were averaged across the 20 repeats for each comparison before calculating the primary performance estimates. Additional metrics included the area under the precision-recall curve (AUC-PR), accuracy, sensitivity, specificity, positive predictive value, negative predictive value, F1 score, Brier score, and log loss. Classification metrics used a prespecified probability threshold of 0.50. Confidence intervals were estimated using 10,000 trial-cluster bootstrap resamples. Incremental performance from adding PRO features was quantified using paired trial-cluster bootstrap differences in AUC-ROC.

Model interpretability was assessed using held-out linear SHapley Additive exPlanations (SHAP) values from 20 repeats of five-fold stratified trial-grouped cross-validation. In each fold, imputation, scaling, encoding, and model fitting were learned from the training data, and the corresponding transformed training data were used as the SHAP background. SHAP values were calculated for held-out observations on the log-odds scale. Transformed components, including one-hot-encoded levels and missingness indicators, were summed within their original feature before mean absolute SHAP values were calculated across held-out observations. SHAP importance therefore represents predictive influence rather than statistical significance or causality.

### Software and Codes Availability

Univariate logistic regression was performed using performed using R 4.4.1 and RStudio version 2022.07.1+554. Endpoint harmonization, machine-learning analyses, bootstrap resampling, and SHAP analyses were performed in Python 3.10.2 using pandas, statsmodels, scikit-learn, and SHAP packages. Final figure formatting was performed using GraphPad Prism 10. The machine-learning code is available at https://github.com/Gracellcc/qol-os-benefit-ml.

### Data Availability

Data used in this study are publicly available. The database and processed dataset for the final analysis dataset is available at https://github.com/Gracellcc/qol-os-benefit-ml.

## Results

### Study and Participants Inclusion

Between 2013 and 2023, 384 oncology trials supporting FDA drug approvals were identified, of which 380 passed initial screening. Among these, 100 trials (26.3%) had publicly available data on OS, at least one surrogate efficacy endpoint, and PROs and were eligible for analysis. These trials included 68,987 participants (36,686 in active arms and 32,301 in control arms) and contributed 103 between-arm comparisons to the modeling analyses. The trial inclusion and exclusion process is shown in **Figure 1**, and study characteristics are summarized in **Table 1**.

**Table 1.** Characteristics of Randomized Clinical Trials Between-Arm Comparisons Included in the Analysis.

|  |  |  |  |
| --- | --- | --- | --- |
| **Overall Survival Benefits** | No<br>33 (32.0%) | Yes<br>70 (68.0%) | Total<br>103 |
| **Drug Class** |  |  |  |
| Immunotherapy | 5 (15.2%) | 26 (37.1%) | 31 (30.1%) |
| Targeted Therapy | 21 (63.6%) | 30 (42.9%) | 51 (49.5%) |
| Immunotherapy plus Targeted Therapy | 1 (3.0%) | 2 (2.9%) | 3 (2.9%) |
| Other (e.g., Chemotherapy, Hormonal) | 6 (18.2%) | 12 (17.1%) | 18 (17.5%) |
| **Disease** |  |  |  |
| Genitourinary (GU) | 6 (18.2%) | 14 (20.0%) | 20 (19.4%) |
| Breast | 6 (18.2%) | 10 (14.3%) | 16 (15.5%) |
| Gastrointestinal (GI) | 4 (12.1%) | 9 (12.9%) | 13 (12.6%) |
| Hematologic malignancies | 4 (12.1%) | 10 (14.3%) | 14 (13.6%) |
| Lung | 5 (15.2%) | 11 (15.7%) | 16 (15.5%) |
| Gynecologic | 3 (9.1%) | 3 (4.3%) | 6 (5.8%) |
| Melanoma | 1 (3.0%) | 8 (11.4%) | 9 (8.7%) |
| Other | 4 (12.1%) | 5 (7.1%) | 9 (8.7%) |
| **Surrogate endpoints benefits** |  |  |  |
| Yes | 27 (81.8%) | 59 (84.3%) | 86 (83.5%) |
| No/Unknown | 6 (18.2%) | 11 (15.7%) | 17 (16.5%) |
| **PRO endpoints benefits** |  |  |  |
| Yes | 11 (33.3%) | 26 (37.1%) | 37 (35.9%) |
| No/Unknown | 22 (66.7%) | 44 (62.9%) | 66 (64.1%) |

### PROs are Underrepresented in Drug Development

Among the 103 between-arm comparisons, 70 (68.0%) demonstrated an OS benefit, 86 (83.5%) showed improvement in surrogate endpoints, and 37 (35.9%) showed PRO improvement. Of the 33 comparisons without an OS benefit, 27 (81.8%) nevertheless showed improvement in surrogate endpoints, suggesting that surrogate endpoint benefit did not consistently translate into OS benefit. In the same group, 11 (33.3%) showed PRO improvement, whereas 22 (66.7%) did not show a clear PRO benefit. Among the 70 comparisons with an OS benefit, 26 (37.1%) showed PRO improvement and 44 (62.9%) did not. Overall, neither surrogate nor PRO endpoints alone clearly distinguished trials with or without OS benefit, supporting evaluation of their combined predictive value (**Figure 2A**).

**Figure 2.**
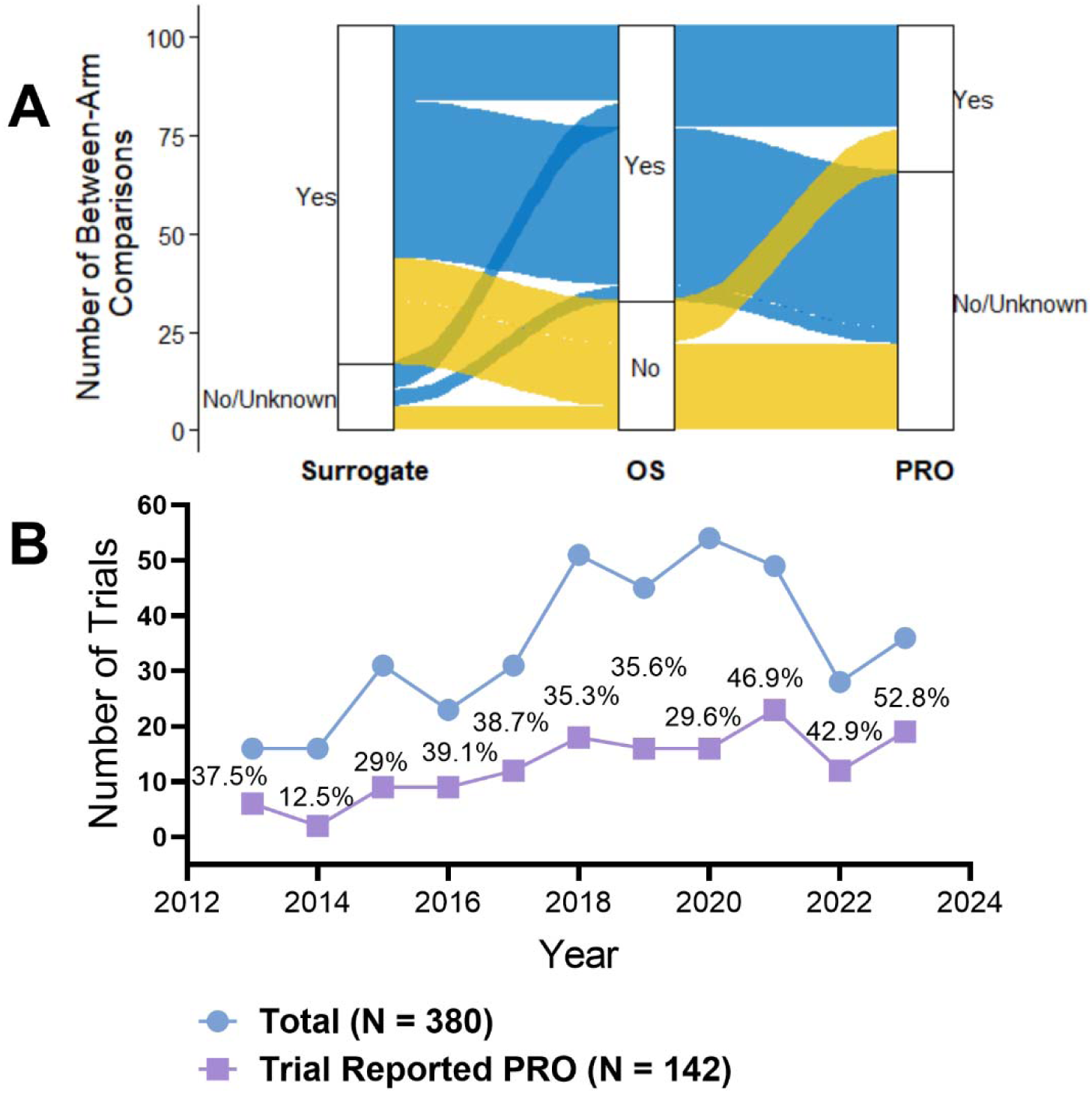
PROs are underrepresented in drug development. (A) Sankey diagram illustrating the overlap among oncology trials demonstrating benefits in OS, surrogate efficacy endpoints (e.g., progression-free survival or overall response rate), and PROs. (B) Numbers and percentages of trials reporting PROs among all approvals in each year.

We also assessed the availability of PRO data in oncology trials over time. Among the 380 trials that passed screening, 142 (37.4%) reported PROs. (**Figure 2B**) The proportion of trials reporting PROs increased over time, consistent with growing efforts by regulators and sponsors to promote the inclusion and reporting of PROs in oncology clinical trials. (15)

### Association Between PRO Endpoints, Surrogate Endpoints, and OS

Univariate logistic regression models were developed to evaluate the associations between PRO endpoints, surrogate endpoints, and OS benefits. The unadjusted odds ratios (ORs) and corresponding 95% confidence intervals (CIs) are presented in **Figure 3**. An OR greater than 1 suggests that patients (or trials) with this characteristic had a higher likelihood of improved OS. Trials with PRO benefits had higher odds of demonstrating OS benefits compared with those without PRO benefits (OR = 1.18 [95% CI: 0.5–2.89]). A similar trend was observed for surrogate endpoints (OR = 1.19 [95% CI: 0.38–3.48]). However, none of these associations reached statistical significance (p > 0.05). These findings suggest that the relationships among OS, surrogate endpoints, and PROs are complex and may require advanced analytical approaches, motivating the use of machine learning to integrate treatment effects, uncertainty, sample size, endpoint characteristics, and cross-domain information.

**Figure 3.**
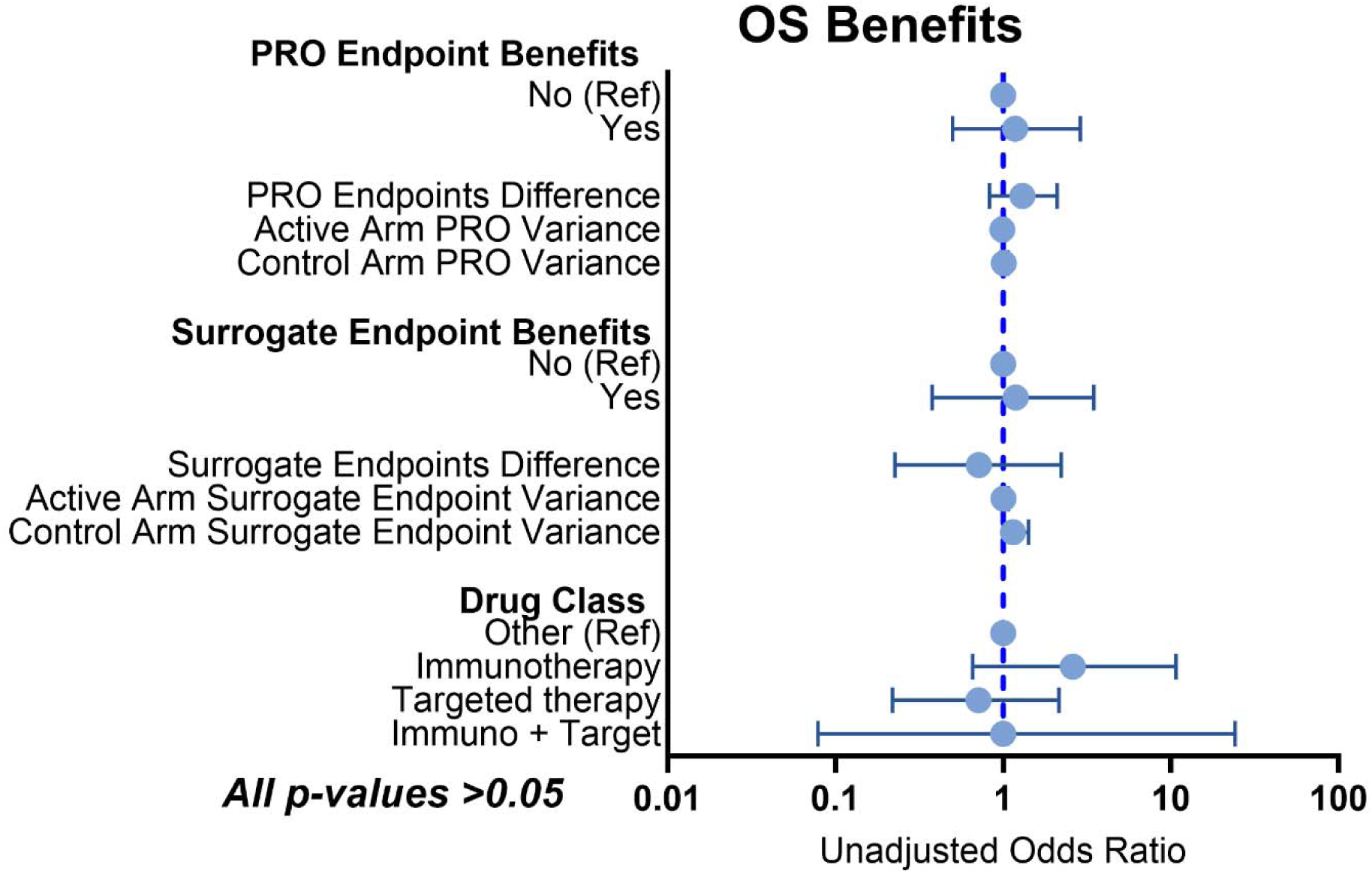
Univariate logistic regression shows unadjusted odds ratios with 95% confidence intervals for patient-reported outcome (PRO) endpoints, surrogate endpoints, and drug class associations with overall survival (OS) benefit. No statistically significant associations were observed, with all p-values > 0.05. Unadjusted odd ratio > 1 indicates a higher likelihood of OS benefit.

### Integrating PROs Improves OS Benefit Prediction via Machine Learning

We then evaluated the performance of machine learning models trained to predict OS benefit. AUC-ROC represents how well the model can discriminate between trials with and without an OS benefit, with higher values indicating better model performance. Regularized logistic regression demonstrated the strongest and most stable performance across the candidate algorithms and was selected as the final model. Based on discrimination, stability, and the sample size, the final model was locked as an L2-regularized logistic regression with C=1. In 20 repeats of five-fold stratified trial-grouped cross-validation, the model using other efficacy endpoints alone achieved an AUC-ROC of 0.668 (95% CI, 0.545–0.785). Adding PRO features increased the AUC-ROC to 0.786 (95% CI, 0.686–0.873), an absolute paired increase of 0.117 (95% CI, −0.003 to 0.237). (**Figure 4A**) The combined model also improved accuracy (0.725 to 0.770), specificity (0.354 to 0.523), precision (0.744 to 0.795), and F1 score (0.816 to 0.839). (**Table 2**) Leave-one-trial-out validation confirmed the added predictive value of PROs: AUC-ROC increased from 0.663 (95% CI, 0.540–0.778) to 0.795 (95% CI, 0.701–0.879), with a paired increase of 0.132 (95% CI, 0.017–0.248). Improvements remained directionally consistent in sensitivity analyses excluding investigator-assigned benefit labels and excluding sample-size-related predictors. Collectively, these results demonstrate that PRO features contributed incremental information beyond conventional efficacy endpoints for internally validated prediction of OS benefit.

**Figure 4.**
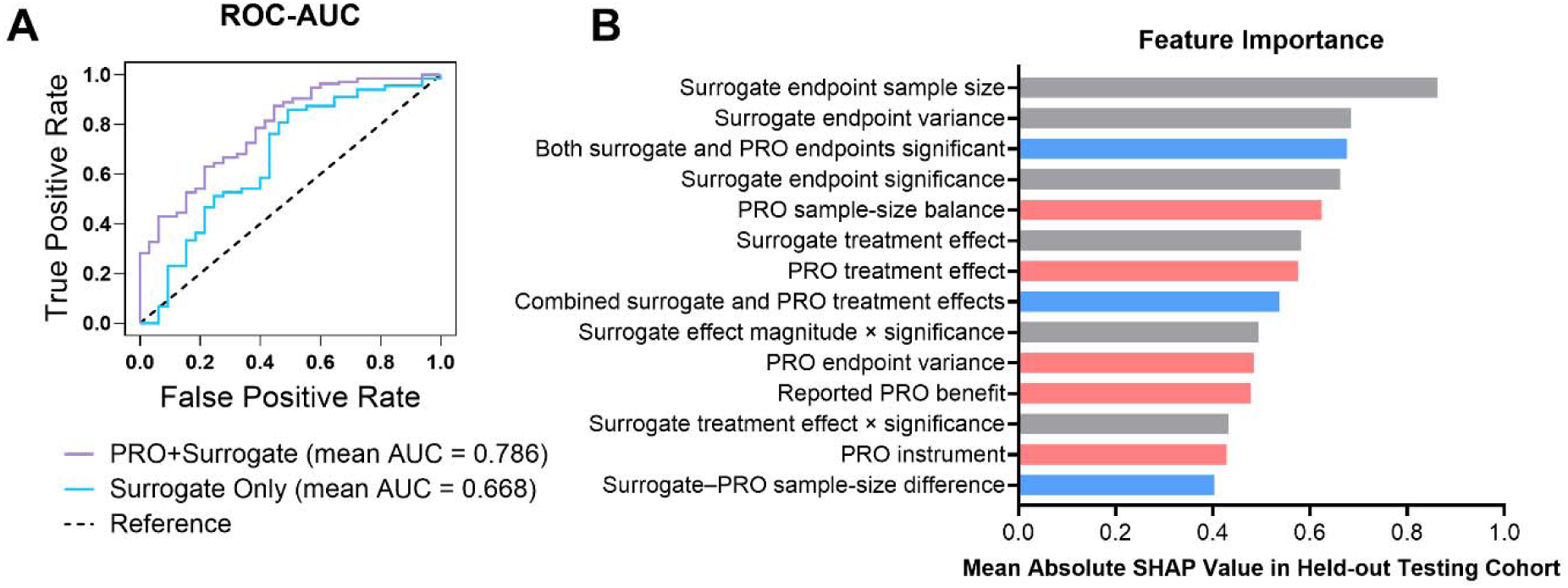
Added value of patient-reported outcomes (PROs) for predicting overall survival (OS) benefit using machine learning. (A) Receiver operating characteristic (ROC) curve for models using surrogate endpoint features alone versus surrogate and PRO features combined. Higher area under the curve (AUC) of ROC indicates better model discrimination between with and without OS benefit. (B) Feature importance based on SHapley Additive exPlanations (SHAP) values for the model incorporating both surrogate and PRO features. Higher SHAP values indicate greater contributions to model predictions. Red bars represent PRO-related features, gray bars surrogate-related features, and blue bars cross-domain features. Only features with SHAP values >0.4 are shown.

**Table 2.**
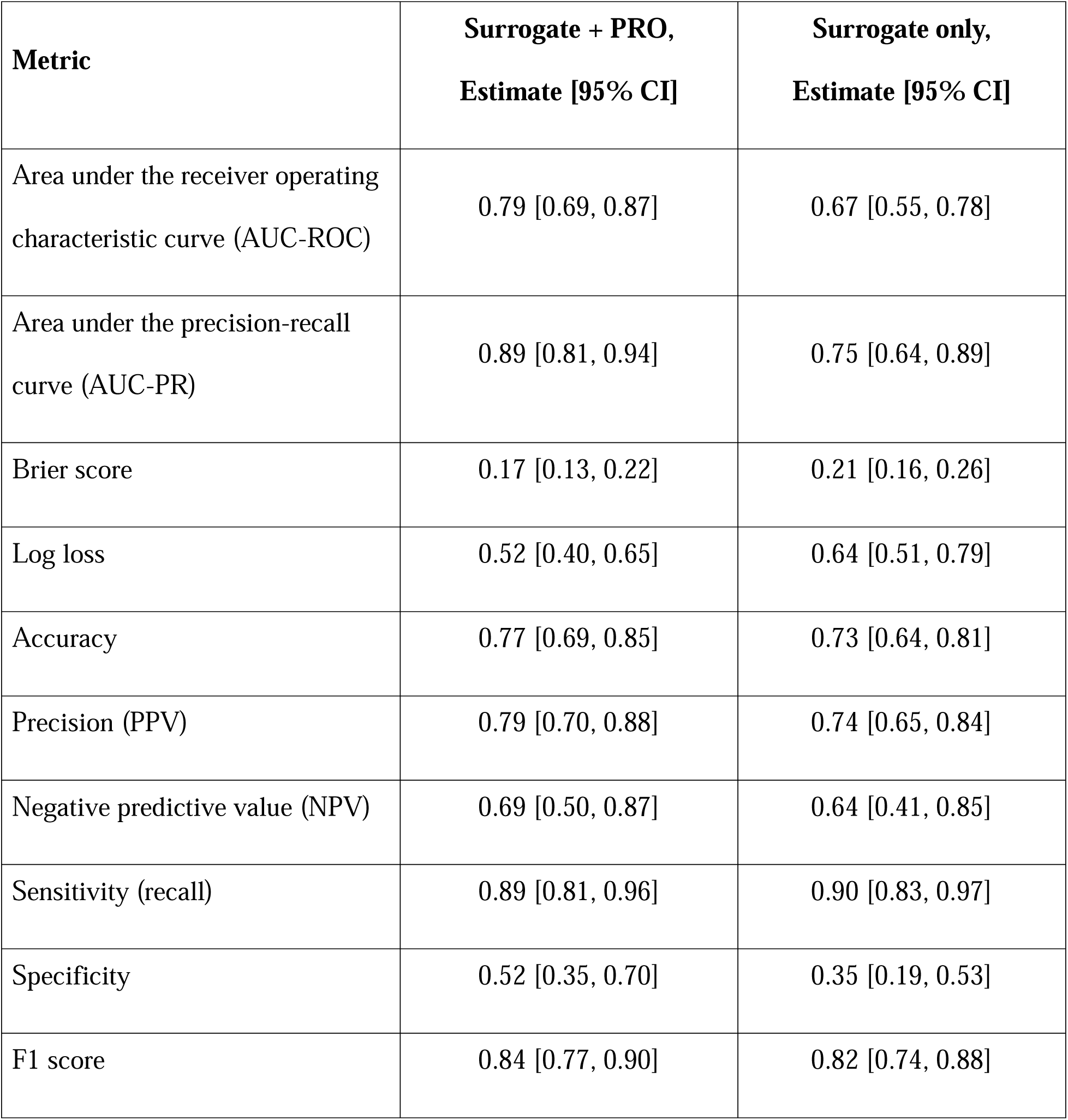
Performance of machine learning models with and without patient-reported outcome (PRO) features.

Held-out SHAP analysis was conducted to identify the contributions from both surrogate and PRO domains (**Figure 4B**). The most influential feature was the log-transformed total sample size for the surrogate efficacy endpoint (mean absolute SHAP, 0.862), followed by its variance (0.684), concurrent significance for the surrogate and PRO endpoints (0.676), surrogate endpoint significance (0.662), and the active-to-control PRO sample-size log ratio in PRO endpoints (0.624). Additional influential PRO features included the direction-adjusted PRO treatment effect (0.575), PRO endpoint variance (0.484), reported PRO benefit status (0.478), and PRO instrument family (0.429). The prominence of multiple PRO-related and cross-domain features supports the incremental contribution of PRO information to model predictions. These SHAP values quantify predictive influence on the log-odds scale and should not be interpreted as causal effects.

## Discussion

This study demonstrates that PROs provide complementary information for predicting OS benefit when integrated with other efficacy endpoints. Using 103 between-arm comparisons from 100 FDA approval-linked oncology trials, the machine learning model showed a substantial improvement in internally validated discrimination after PRO features were added. The improvement was consistent across repeated trial-grouped cross-validation, leave-one-trial-out validation, and sensitivity analyses, supporting the robustness of the incremental PRO signal.

We first examined the associations of PROs and surrogate endpoints with OS benefit using univariate logistic regression, but no statistically significant relationships were identified, likely because this approach does not capture complex interactions among endpoint characteristics. (16) In contrast, machine learning can integrate multiple complementary features, including treatment effect magnitude, precision, sample size, endpoint type, and concordance between surrogate and PRO domains, while controlling model complexity and maintaining trial-level separation during validation. (17,18) These findings highlight the potential of machine learning models to integrate multidimensional clinical trial data and improve prediction of treatment benefit, supporting its future use as a quantitative tool for trial evaluation and drug development.(19)

Although formal validation of surrogacy requires patient-level data within specific disease settings, the present study addresses a complementary question: whether trial-level PRO information improves prediction of OS benefit beyond other efficacy endpoints. Our findings have demonstrated that adding PROs provided additional predictive value. Consistent with this observation, our previously published study using individual participant-level PRO and PFS data in breast cancer found that adding PROs to PFS improved prediction of OS using machine learning models. (14) Together, these findings support further evaluation of integrated surrogate–PRO approaches for improving prediction of long-term clinical benefit in oncology trials.

This study has several limitations. The analysis was restricted to trials with available OS, surrogate efficacy endpoints, and PRO data, resulting in a modest sample size and potential reporting-selection bias. PRO measures and reporting conventions varied across trials; direction harmonization and scale-invariant engineered features reduced, but could not eliminate, this heterogeneity. Model performance was evaluated using rigorous internal validation, including trial-grouped repeated cross-validation and leave-one-trial-out validation, but independent external validation remains necessary to quantify transportability across cancer types, treatment classes, and future approvals. (20)

In summary, integrating PROs with conventional surrogate efficacy endpoints improved trial-level prediction of OS benefit in a rigorously validated regularized machine-learning framework. PRO-related features made substantive contributions to model predictions, demonstrating that patient-reported information captures clinically relevant signals not fully represented by surrogate efficacy endpoints alone. With external validation, this framework could strengthen early evaluation of long-term clinical benefit and support more comprehensive, patient-centered evidence generation in oncology drug development.

## Notes

**Conflicts of Interest:** B.M reports personal fees from Novartis outside this submitted work. E.B. receives institutional research funding from the National Cancer Institute and from the Patient-Centered Outcomes Research Institute, and personal fees for scientific advising from Research Triangle Institute, Thyme Care, N-Power Medicine, Resilience Health, Canopy Care, Savor Health, and Navigating Cancer.

### Competing Interest Statement

B.M reports personal fees from Novartis outside this submitted work. E.B. receives institutional research funding from the National Cancer Institute and from the Patient-Centered Outcomes Research Institute, and personal fees for scientific advising from Research Triangle Institute, Thyme Care, N-Power Medicine, Resilience Health, Canopy Care, Savor Health, and Navigating Cancer.

### Author Declarations

The study used ONLY openly available human data that were originally located at FDA oncology New Drug Registry.

### Summary of Updates

This version of the manuscript has been revised to improve the accuracy and reproducibility of the underlying database. A newly assembled team collected the data again, corrected several errors identified in the previous version, and cross checked every record. We then repeated all analyses using the corrected and verified database and updated the manuscript accordingly. The methods and main conclusions remain unchanged. These revisions strengthen confidence in the findings by ensuring that the reported results are based on a thoroughly checked database that supports reproducible analyses.

